# Endogenous glucagon-like peptide 1 diminishes prandial glucose counterregulatory response to hypoglycemia after gastric bypass surgery

**DOI:** 10.1101/2023.09.20.23295840

**Authors:** Henri Honka, Amalia Gastaldelli, Samantha Pezzica, Richard Peterson, Ralph DeFronzo, Marzieh Salehi

**Affiliations:** Division of Diabetes, University of Texas Health Science Center, San Antonio, TX; Cardiometabolic Risk Unit, Institute of Clinical Physiology-National Research Council, Pisa, Italy; Department of Surgery, University of Texas Health Science Center, San Antonio, TX; South Texas Veteran Health Care System, Audie Murphy Hospital, San Antonio, TX

**Keywords:** gastric bypass surgery, sleeve gastrectomy, hypoglycemia, counterregulatory response, glucagon-like peptide 1

## Abstract

We have previously shown that prandial endogenous glucose production (*EGP*) during insulin-induced hypoglycemia is smaller in non-diabetic subjects with gastric bypass (GB), where prandial glucagon-like peptide 1 (GLP-1) concentrations are 5-10 times higher than those in non-operated controls. Here, we sought to determine the effect of *endogenous* GLP-1 on prandial counterregulatory response to hypoglycemia after GB. Glucose fluxes, and islet-cell and gut hormone responses before and after mixed-meal ingestion were compared during a hyperinsulinemic hypoglycemic (**∽**3.2 mmol/l) clamp with and without a GLP-1 receptor (GLP-1R) antagonist exendin-(9-39) (Ex-9) in non-diabetic subjects with prior GB compared to matched subjects with SG and non-surgical controls. In this setting, GLP-1R blockade had no effect on insulin secretion or insulin action, whereas prandial glucagon was enhanced in all 3 groups. Ex-9 infusion raised prandial *EGP* response to hypoglycemia in every GB subject but had no consistent effects on EGP among subjects with SG or non-operated controls (*P* < 0.05 for interaction). These results indicate that impaired post-meal glucose counterregulatory response to hypoglycemia after GB is partly mediated by endogenous GLP-1, highlighting a novel mechanism of action of GLP-1R antagonists for the treatment of prandial hypoglycemia in this population.

## INTRODUCTION

Glucagon-like peptide 1 (GLP-1) plays a key regulatory role in prandial glucose metabolism, primarily by increasing insulin secretion and suppressing glucagon secretion [1] in a glucose-dependent fashion [2]. While by reducing glucagon-to-insulin ratio, GLP-1 can potentially reduce endogenous glucose production (*EGP*) [3]. Exogenous GLP-1 infusion has also been shown to diminish *EGP* by ∽20 % under euglycemic conditions [4] and by ∽60 % in the hyperglycemic state [5], suggestive of extrapancreatic GLP-1 effect.

In non-diabetic humans without history of gastrointestinal tract-altering surgery, the recovery from hypoglycemia is brought about by early inhibition of insulin secretion in response to glycemic decline, which lowers glucose utilization, followed by stimulation of counterregulatory hormones, mainly glucagon, which raise glucose production [6-8]. We have recently reported that prandial *EGP* response to insulin-induced hypoglycemia is diminished in non-diabetic patients with prior history of bariatric surgery, particularly in those with gastric bypass (GB) compared to matched non-operated controls despite higher prandial glucagon concentrations, indicating resistance to glucagon [9].

It is well recognized that following GB, meal ingestion increases prandial GLP-1 secretion by 5-10 fold [10-13], particularly in those who develop the late-complication of hyperinsulinemic hypoglycemic syndrome [14-16]. Further, we and others have shown that blocking GLP-1 receptor (GLP-1R) by exendin-(9-39) (Ex-9) infusion diminishes insulin secretory response to meal ingestion in subjects after GB [17-20] and corrects postprandial hypoglycemia in patients with recurrent hypoglycemia after GB [16, 21]. While the effect of endogenous GLP-1 on *EGP* response to hypoglycemia in humans is largely unknown, it is plausible that enhanced prandial GLP-1 secretion after GB [10-12] also disturbs the glucose counterregulatory response to hypoglycemia after meal ingestion.

Enhanced prandial GLP-1 secretion [13, 22] and GLP-1-induced insulin response [13] after GB is shared with SG but in much smaller magnitude. Therefore, we hypothesized that endogenous GLP-1 contributes to blunted prandial *EGP* response to hypoglycemia after GB. To test this hypothesis, we examined post-meal glucose fluxes, and islet-cell and gut hormone responses during hyperinsulinemic hypoglycemic clamp with and without blocking GLP-1R by continuous infusion of Ex-9 in non-diabetic subjects with prior history of GB compared to BMI- and age-matched subjects with SG, as well as non-surgical controls.

## RESULTS

### Subject characteristics (Table 1)

The subjects in GB, SG and CN groups had similar BMI, waist circumference, body fat mass and lean mass, age, sex and HbA_1c_. Time since surgery and post-operative weight loss were similar among the two surgical groups.

**Table 1.**
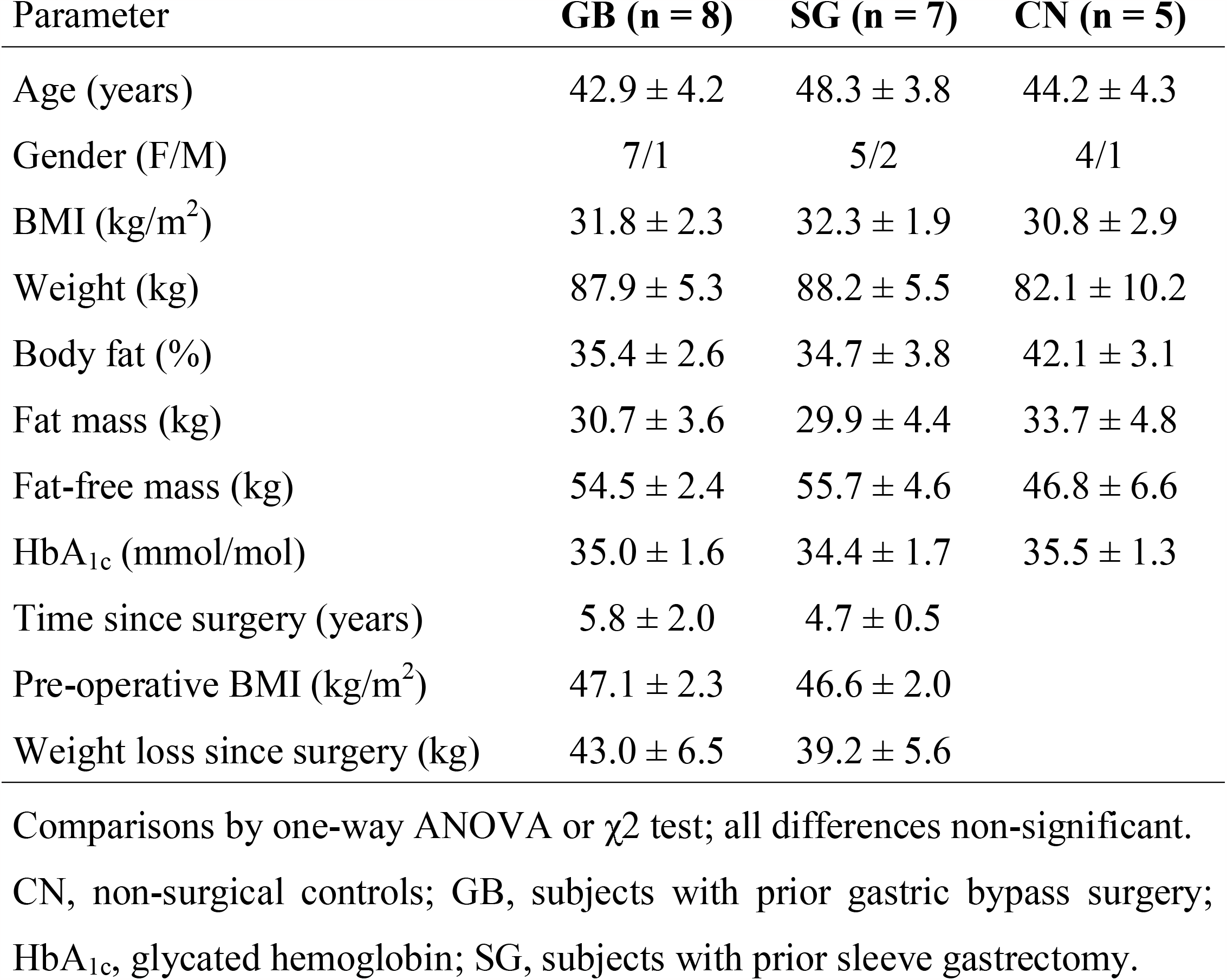
Subject characteristics.

### Physiologic responses before and after meal ingestion during hyperinsulinemic hypoglycemic clamp with and without blocking GLP-1R

Portions of glucose, insulin and glucagon data during the saline studies previously were reported as part of an investigation that focused on the insulin clearance and glucose counterregulatory response to hypoglycemia [9, 23]. The effect of endogenous GLP-1 on glucose fluxes and hormonal response are reported in the current manuscript.

### Glucose and GIR (Fig. 1)

Fasting plasma glucose concentration was lower in SG-treated subjects than in GB-treated or CN subjects (*P* < 0.05), and lower during Ex-9 studies compared to saline studies (*P* < 0.05) (**Table 2**). By the steady-state period (110-120 min), plasma glucose was similarly reduced to the target value of **∽**3.2 mmol/l during studies with and without Ex-9 infusion in all 3 groups. Glucose infusion rates at steady-state period were similar among the groups.

**Table 2.**
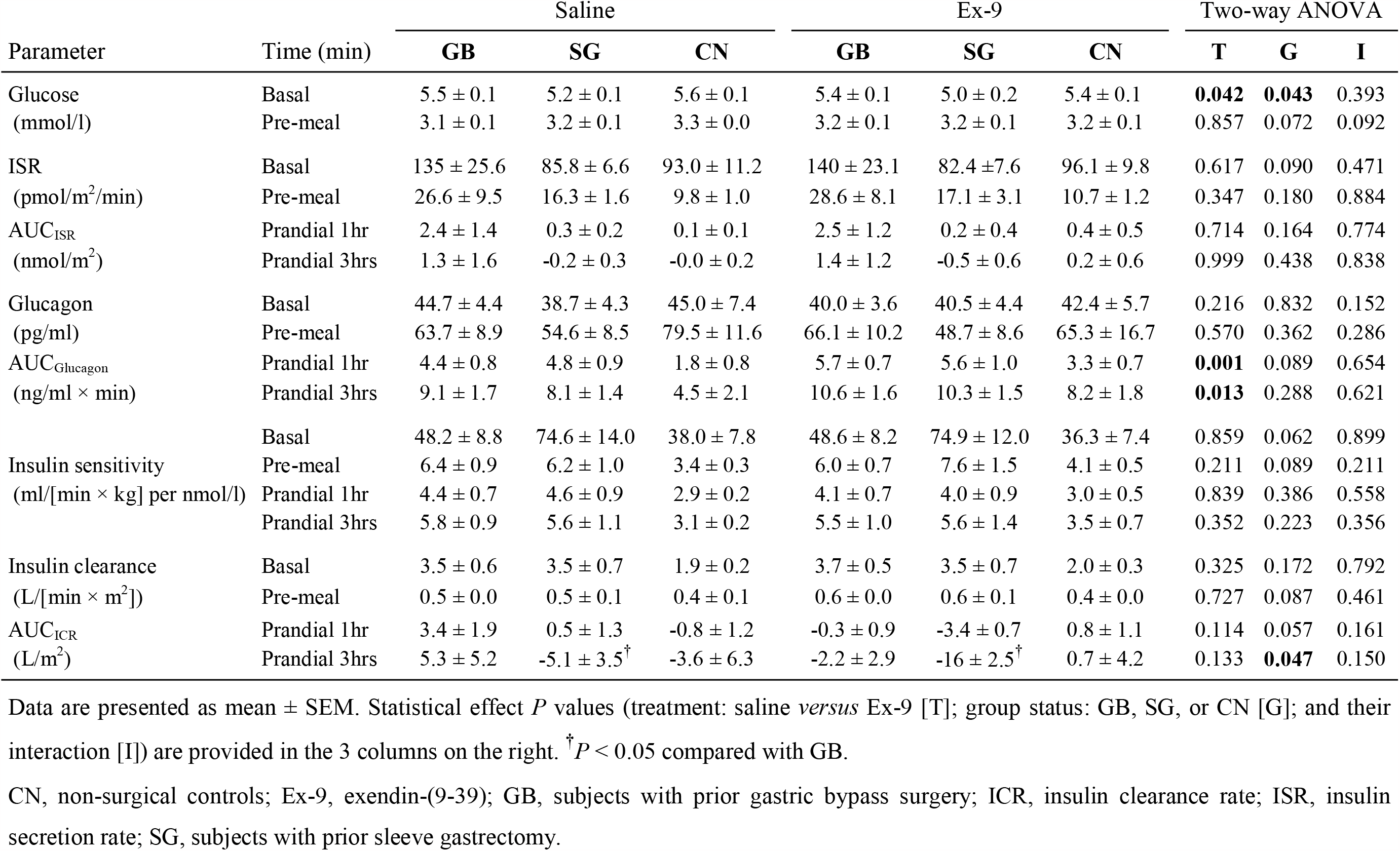
Glucose, islet cell and insulin kinetic responses to insulin-induced hypoglycemia and mixed-meal test during saline and exendin-(9-39) infusion in GB, SG and CN groups.

| Parameter | Time (min) | Saline |  |  | Ex-9 |  |  | Two-way ANOVA |  |  |
| --- | --- | --- | --- | --- | --- | --- | --- | --- | --- | --- |
|  |  | GB | SG | CN | GB | SG | CN | T | G | I |
| Glucose (mmol/l) | Basal | 5.5 ± 0.1 | 5.2 ± 0.1 | 5.6 ± 0.1 | 5.4 ± 0.1 | 5.0 ± 0.2 | 5.4 ± 0.1 | **0.042** | **0.043** | 0.393 |
|  | Pre-meal | 3.1 ± 0.1 | 3.2 ± 0.1 | 3.3 ± 0.0 | 3.2 ± 0.1 | 3.2 ± 0.1 | 3.2 ± 0.1 | 0.857 | 0.072 | 0.092 |
| ISR (pmol/m<sup>2</sup>/min) | Basal | 135 ± 25.6 | 85.8 ± 6.6 | 93.0 ± 11.2 | 140 ± 23.1 | 82.4 ± 7.6 | 96.1 ± 9.8 | 0.617 | 0.090 | 0.471 |
|  | Pre-meal | 26.6 ± 9.5 | 16.3 ± 1.6 | 9.8 ± 1.0 | 28.6 ± 8.1 | 17.1 ± 3.1 | 10.7 ± 1.2 | 0.347 | 0.180 | 0.884 |
| AUC<sub>ISR</sub> (nmol/m<sup>2</sup>) | Prandial 1hr | 2.4 ± 1.4 | 0.3 ± 0.2 | 0.1 ± 0.1 | 2.5 ± 1.2 | 0.2 ± 0.4 | 0.4 ± 0.5 | 0.714 | 0.164 | 0.774 |
|  | Prandial 3hrs | 1.3 ± 1.6 | -0.2 ± 0.3 | -0.0 ± 0.2 | 1.4 ± 1.2 | -0.5 ± 0.6 | 0.2 ± 0.6 | 0.999 | 0.438 | 0.838 |
| Glucagon (pg/ml) | Basal | 44.7 ± 4.4 | 38.7 ± 4.3 | 45.0 ± 7.4 | 40.0 ± 3.6 | 40.5 ± 4.4 | 42.4 ± 5.7 | 0.216 | 0.832 | 0.152 |
|  | Pre-meal | 63.7 ± 8.9 | 54.6 ± 8.5 | 79.5 ± 11.6 | 66.1 ± 10.2 | 48.7 ± 8.6 | 65.3 ± 16.7 | 0.570 | 0.362 | 0.286 |
| AUC<sub>Glucagon</sub> (ng/ml × min) | Prandial 1hr | 4.4 ± 0.8 | 4.8 ± 0.9 | 1.8 ± 0.8 | 5.7 ± 0.7 | 5.6 ± 1.0 | 3.3 ± 0.7 | **0.001** | 0.089 | 0.654 |
|  | Prandial 3hrs | 9.1 ± 1.7 | 8.1 ± 1.4 | 4.5 ± 2.1 | 10.6 ± 1.6 | 10.3 ± 1.5 | 8.2 ± 1.8 | **0.013** | 0.288 | 0.621 |
| Insulin sensitivity (ml/[min × kg] per nmol/l) | Basal | 48.2 ± 8.8 | 74.6 ± 14.0 | 38.0 ± 7.8 | 48.6 ± 8.2 | 74.9 ± 12.0 | 36.3 ± 7.4 | 0.859 | 0.062 | 0.899 |
|  | Pre-meal | 6.4 ± 0.9 | 6.2 ± 1.0 | 3.4 ± 0.3 | 6.0 ± 0.7 | 7.6 ± 1.5 | 4.1 ± 0.5 | 0.211 | 0.089 | 0.211 |
|  | Prandial 1hr | 4.4 ± 0.7 | 4.6 ± 0.9 | 2.9 ± 0.2 | 4.1 ± 0.7 | 4.0 ± 0.9 | 3.0 ± 0.5 | 0.839 | 0.386 | 0.558 |
|  | Prandial 3hrs | 5.8 ± 0.9 | 5.6 ± 1.1 | 3.1 ± 0.2 | 5.5 ± 1.0 | 5.6 ± 1.4 | 3.5 ± 0.7 | 0.352 | 0.223 | 0.356 |
| Insulin clearance (L/[min × m<sup>2</sup>]) | Basal | 3.5 ± 0.6 | 3.5 ± 0.7 | 1.9 ± 0.2 | 3.7 ± 0.5 | 3.5 ± 0.7 | 2.0 ± 0.3 | 0.325 | 0.172 | 0.792 |
|  | Pre-meal | 0.5 ± 0.0 | 0.5 ± 0.1 | 0.4 ± 0.1 | 0.6 ± 0.0 | 0.6 ± 0.1 | 0.4 ± 0.0 | 0.727 | 0.087 | 0.461 |
| AUC<sub>ICR</sub> (L/m<sup>2</sup>) | Prandial 1hr | 3.4 ± 1.9 | 0.5 ± 1.3 | -0.8 ± 1.2 | -0.3 ± 0.9 | -3.4 ± 0.7 | 0.8 ± 1.1 | 0.114 | 0.057 | 0.161 |
|  | Prandial 3hrs | 5.3 ± 5.2 | -5.1 ± 3.5<sup>†</sup> | -3.6 ± 6.3 | -2.2 ± 2.9 | -16 ± 2.5<sup>†</sup> | 0.7 ± 4.2 | 0.133 | **0.047** | 0.150 |

Data are presented as mean ± SEM. Statistical effect *P* values (treatment: saline *versus* Ex-9 [T]; group status: GB, SG, or CN [G]; and their interaction [I]) are provided in the 3 columns on the right. <sup>†</sup>*P* < 0.05 compared with GB.
CN, non-surgical controls; Ex-9, exendin-(9-39); GB, subjects with prior gastric bypass surgery; ICR, insulin clearance rate; ISR, insulin secretion rate; SG, subjects with prior sleeve gastrectomy.

**Figure 1.**
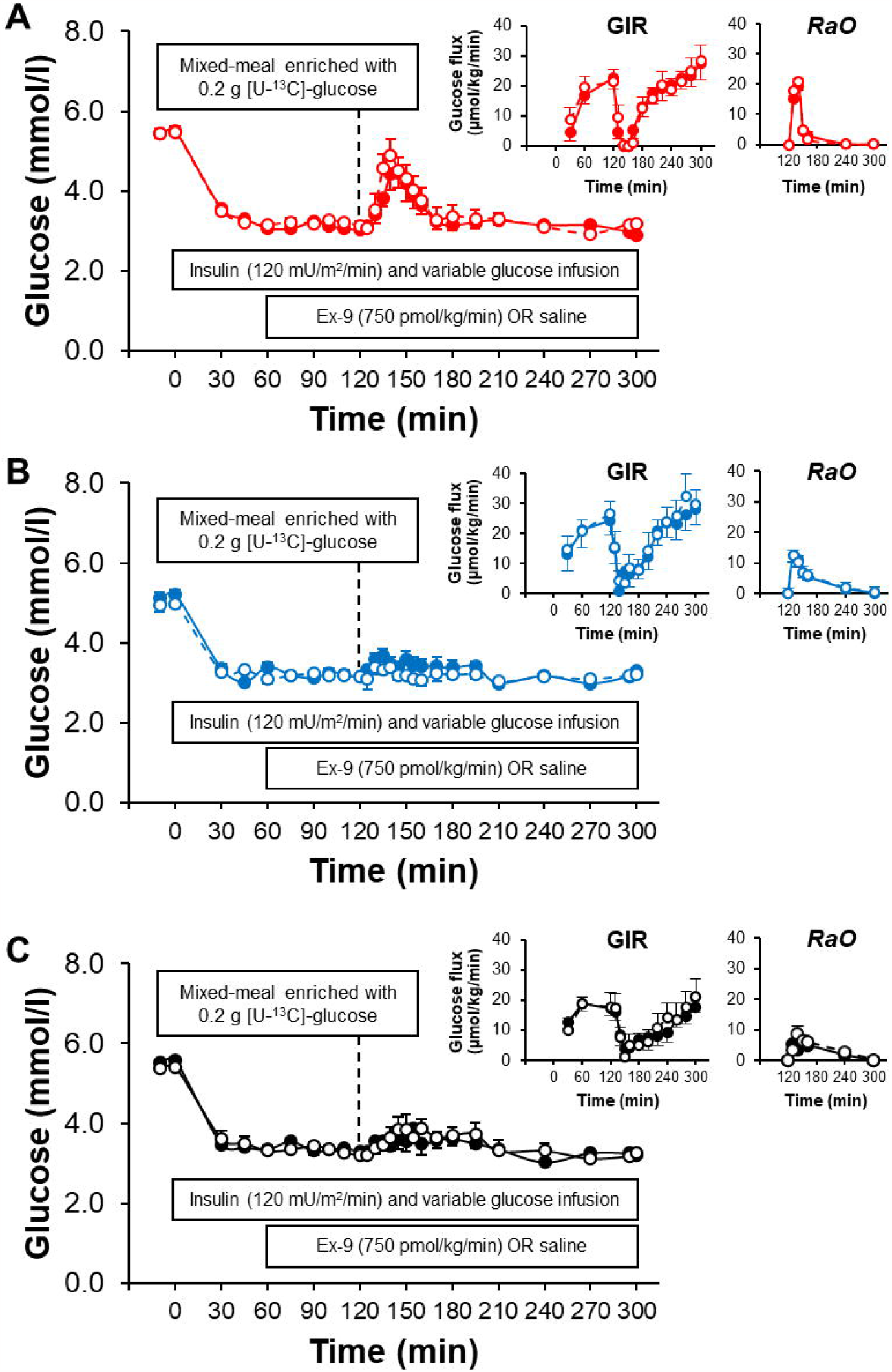
Plasma glucose during hyperinsulinemic hypoglycemic clamp with (dashed line and bar) and without GLP-1R blockade (solid line and bar) in subjects with prior GB (A), SG (B) and non-surgical controls (C). Corresponding rates of glucose infusion (GIR) and ingested oral glucose appearance (RaO) are shown as insets.

To maintain plasma glucose at target levels after meal ingestion, GIR was reduced by ∽90 % in all groups alike (**Fig. 1A-C insets**). Despite a complete discontinuation of the glucose infusion, the average glucose levels were above the target glucose of ∽3.2 mmol/l in 3 GB, 1 SG and 1 CN subject. The average coefficient of variation (CV) of glycemic concentration for each study from 60-300 minutes was higher in GB-treated subjects *versus* SG-treated and controls (16 ± 1.7 % *versus* 11 ± 1.3 and 8.6 ± 1.5 %; *P* < 0.01). However, the average CV of glycemic levels from 60-300 minutes from studies with and without Ex-9 in each subject was 3.1 ± 1.0, 4.9 ± 1.2 and 3.1 ± 0.9 % for GB, SG, and CN, respectively, indicating that glycemic concentrations between the two studies in each subject were identical. The amount of glucose infused to maintain the plasma glucose concentration constant at steady-state or post-meal conditions was unaffected by GLP-1R blockade (**Fig. 1A-C insets**).

### Incretin concentrations (Fig. 2)

Fasting and steady-state incretin concentrations were similar among the groups and between studies with and without Ex-9. Post-meal GLP-1 concentrations over the first 60 minutes of meal study was higher in GB-treated compared with SG or CN (*P* < 0.01). Blockade of GLP-1R similarly increased prandial concentrations of GLP-1 in all 3 groups (**Fig. 2A-C**), consistent with previous reports [16]. Prandial GIP values were similar among the 3 groups and between studies with Ex-9 and saline (**Fig. 2D-F**).

**Figure 2.**
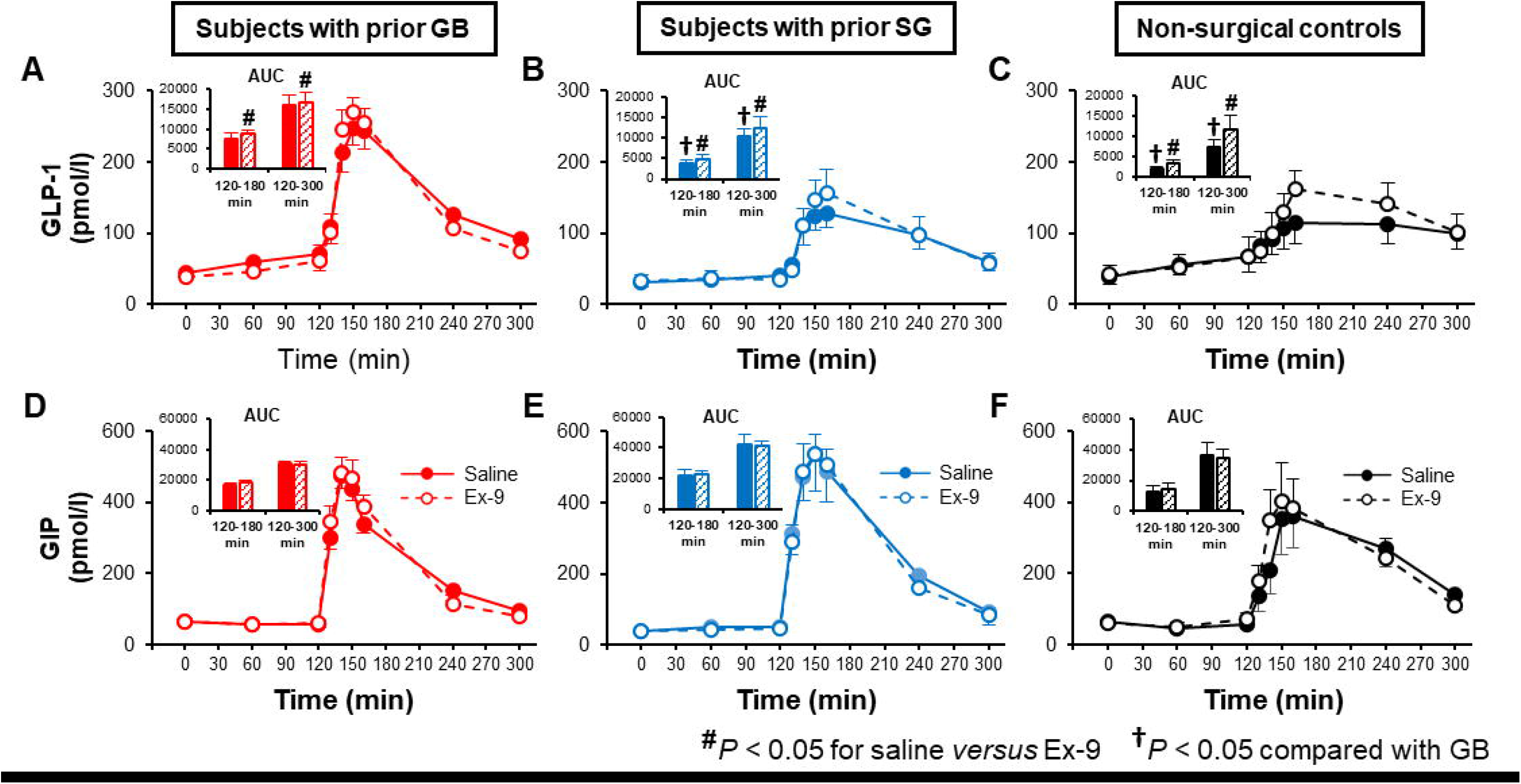
Plasma GLP-1 (A-C) and GIP (D-F) responses to a mixed-meal ingestion during hyperinsulinemic hypoglycemic clamp with (dashed line and bar) and without GLP-1R blockade (solid line and bar) in subjects with prior GB, SG and non-surgical controls. Corresponding AUCs from 120 to 180 min and 120 to 300 min are shown as insets.

### Glucose fluxes (Fig. 3)

After an overnight fast, total glucose utilization (*Rd*) equals endogenous glucose production (*EGP*) and was similar among the three groups and between studies with and without Ex-9 infusion (saline: 9.2 ± 0.4, 9.2 ± 0.2 and 9.3 ± 0.4 μmol/kg/min; Ex-9: 8.9 ± 0.3, 9.7 ± 0.2 and 8.9 ± 0.4 μmol/kg/min for GB, SG and CN, respectively). During the hypoglycemic clamp before meal ingestion, *Rd* progressively increased (**Fig. 3A-C**) whereas *EGP* was suppressed in the first 60 minutes (by ∽90 %) in all 3 groups but rose thereafter due to prolonged hypoglycemia, as previously reported [9] (**Fig 3D-F**). Neither *Rd* nor *EGP* at steady-state before meal consumption were affected by GLP-1R blockade.

**Figure 3.**
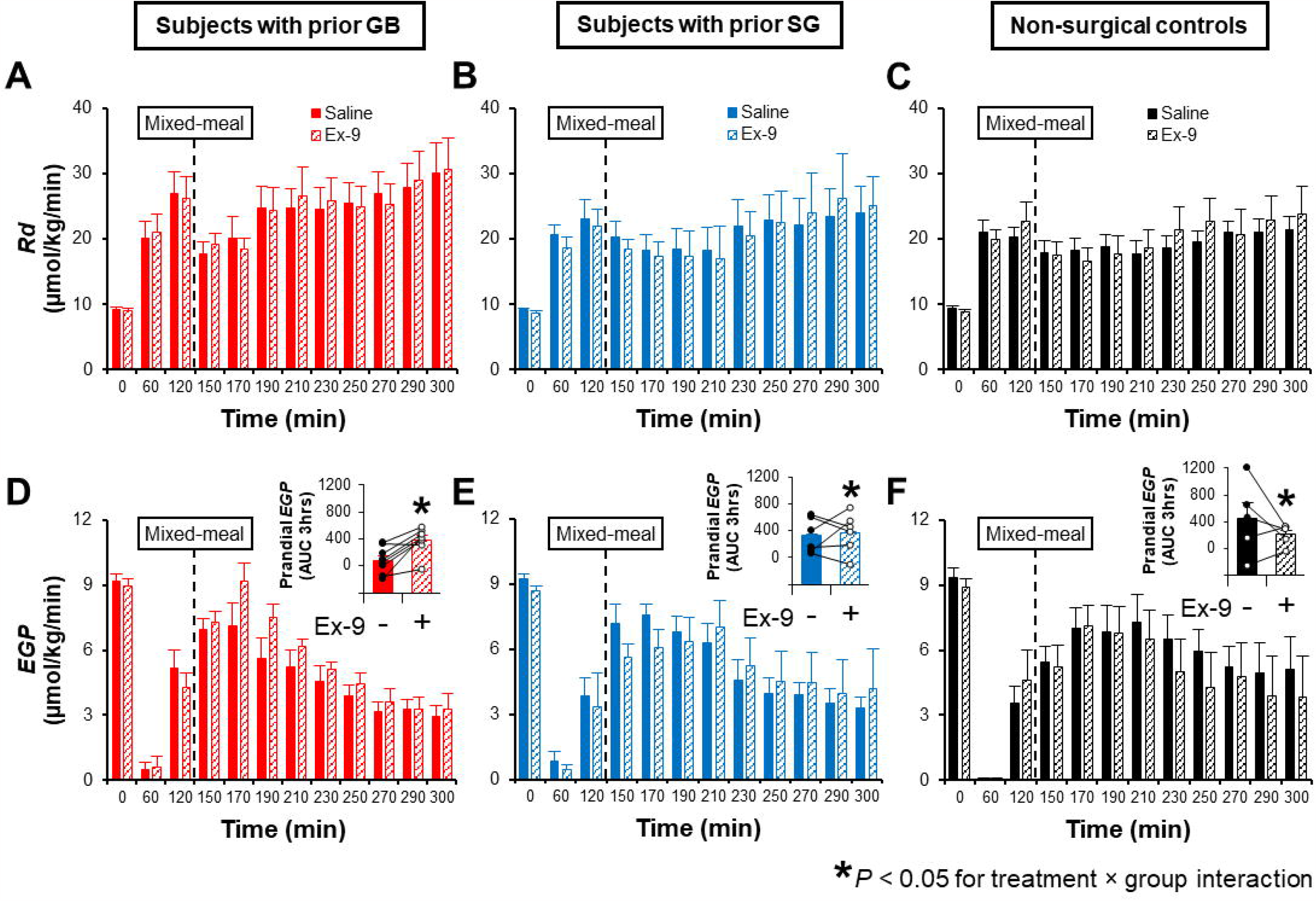
Rates of glucose disappearance (A-C) and endogenous glucose production (D-E) before and after mixed-meal ingestion during hyperinsulinemic hypoglycemic clamp with (dashed bar) and without GLP-1R blockade (solid bar) in subjects with prior GB, SG and non-surgical controls. A differential effect of GLP-1R blockade on AUC *EGP*_3hrs_ (shown as insets) was observed among the groups.

Temporal trends in oral glucose appearance (*RaO*) into the circulation were in line with accelerated gastric emptying by bariatric surgery as AUC *RaO*_1hr_ was higher in GB than SG and in SG compared to CN subjects (*P* < 0.001) (**Fig. 1A-C insets**). However, the AUC *RaO*_3hr_ was similar among the groups (NS). Following meal ingestion total glucose utilization (*Rd*) was similar among the 3 groups (**Fig. 3A-C**). Meal ingestion raised *EGP* levels in the first hour of absorptive period in all 3 groups (**Fig. 3D-F**). Blockade of GLP-1R had no effect on *RaO* or *Rd* after meal ingestion. However, Ex-9 infusion increased prandial *EGP* in every GB subject whereas the *EGP* effect of GLP-1R blockade in SG or CN was significantly inconsistent (*P* < 0.05 for interaction) (**Fig. 3D-F insets**).

Among GB-treated subjects, those with an average post-meal glucose above the target range had a lower *Rd* (**Supplementary Fig. 1A**) compared to those whose glucose remained at the target, despite lack of differences in *RaO* and *EGP* values among the two groups (**Supplementary Fig. 1B-C**).

### Beta- and alpha-cell secretory response (Fig. 4)

Insulin concentrations and ISR were similar at baseline among the groups and between the two studies of Ex-9 or saline infusion (**Fig. 4A-F**). As previously reported [24], the relative reductions in ISR from baseline to steady-state (110-120 min) were attenuated to a larger extent in CN compared to surgical groups (by 82, 81 and 89 % in GB, SG and CN, respectively, *P* < 0.01), but were unaffected by Ex-9 infusion. Fasting plasma glucagon concentrations were similar among the 3 groups and between the two studies, rose slightly during hypoglycemic clamp prior to meal ingestion (**Fig. 4G-I**), but remained unaffected by GLP-1R blockade.

**Figure 4.**
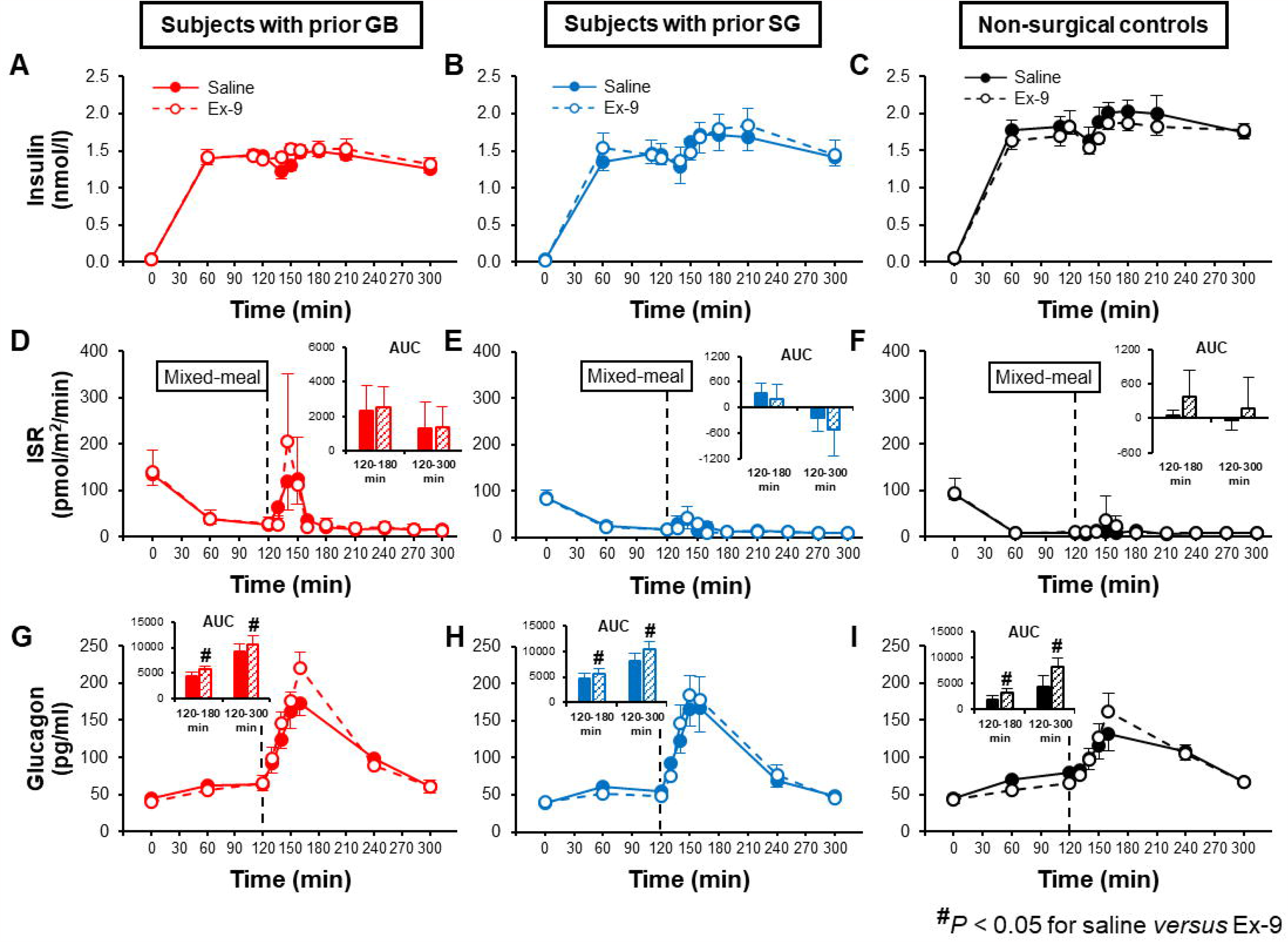
Plasma concentrations of insulin (A-C), ISR (D-F) and glucagon (G-I) before and after mixed-meal ingestion during hyperinsulinemic hypoglycemic clamp with (dashed line and bar) and without GLP-1R blockade (solid line and bar) in subjects with prior history of GB, SG and non-surgical controls. Corresponding AUCs from 120 to 180 min and 120 to 300 min are shown in insets.

During the meal study ISR excursion mimicked that of plasma glucose, with an early transient rise in GB, although prandial AUC 3hr for ISR among the 3 groups did not reach statistical significance (**Fig. 4D-F**). Meal ingestion enhanced plasma glucagon concentrations in all 3 groups (**Fig. 4G-I**), but the levels in the first hour of meal ingestion tended to be higher in surgical than CN subjects (*P* = 0.09).

There was no difference among GB subjects with and without glycemic deviation from target in insulin or glucagon concentrations (**Supplementary Fig. 1D**).

Under hypoglycemic conditions, prandial ISR were unaffected by Ex-9 infusion whereas GLP-1R blockade increased plasma glucagon concentrations in all 3 groups (*P* < 0.05) (**Table 2, Fig. 4G-I**).

### Insulin action and clearance (Table 2)

Whole-body insulin sensitivity (*Rd*/glucose/insulin) at baseline or during steady-state period were similar among the groups (**Table 2**). Basal insulin clearance did not differ among the subjects and was similarly reduced (by ∽80 %) when the plasma insulin concentrations were increased to steady-state levels during the clamp in all 3 groups (**Table 2**). Neither insulin sensitivity nor insulin clearance before meal ingestion were affected by GLP-1R blockade.

Whole-body insulin action in the fed state was similar among the 3 groups and between the two studies with and without Ex-9 infusion (**Table 2**). Prandial insulin clearance (AUC ICR_3hr_) was higher in GB compared with SG (*P* < 0.05) but was unchanged by blocking GLP-1R.

### Heart rate (HR) and blood pressure (BP) (Supplementary Table 1)

At baseline HR and BP were similar among the groups. In response to hypoglycemia HR increased, SBP was unchanged and DBP decreased in all groups alike (**Supplementary Table 1**). Meal ingestion increased HR above pre-meal values by 9 ± 5%, 6 ± 1% and –2 ± 1% in GB, SG and CN, respectively (*P* < 0.05) without any additional effect on BP. During hypoglycemic clamp, blocking the GLP-1R had no effect on HR and BP responses in any of the 3 groups.

## DISCUSSION

The beneficial role of GLP-1 in mediating improved glucose tolerance after gastric bypass surgery by stimulating insulin secretion can be complicated by the development of hyperinsulinemic hypoglycemic syndrome in susceptible subjects after this procedure, where prandial GLP-1 secretion and insulinotropic effect of GLP-1 are exaggerated [25]. In fact, our previous work has shown that blocking GLP-1R corrects prandial hypoglycemia and reduces beta-cell secretion in affected patients [16]. In present study, we add to this knowledge by reporting that endogenous GLP-1 also contributes to the blunted prandial *EGP* response to insulin-induced hypoglycemia in GB-treated subjects without any consistent effect in SG subjects or non-operated controls (**Fig. 3D-F)**. The effect of GLP-1R blockade on counterregulatory response to hypoglycemia is solely observed in prandial condition and not fasting (albeit hyperinsulinemic) state. We also observed that under hypoglycemic condition after meal ingestion, insulinotropic action of GLP-1 is muted while glucagon suppressing effect is preserved regardless of bariatric surgery status. This finding demonstrates a differential glucose-dependency of *endogenous* GLP-1 action on pancreatic beta-cells *versus* alpha-cells in the fed state. This observation makes a stronger case for the use of GLP-1R blocker for treatment of hypoglycemia in this population.

The physiological mechanisms that prevent or correct hypoglycemia are altered after gastric bypass surgery. In this study we demonstrate that the *EGP* response to insulin-induced hypoglycemia in the fed condition is blunted after GB (**Fig. 3D**) [9]. We also demonstrate that endogenous GLP-1 accounts for this impaired post-meal *EGP* response during hypoglycemic clamp after GB (**Fig. 3D**). We cannot discern whether variations in prandial *EGP* response to hypoglycemia among the GB *versus* SG or non-surgical groups is caused by a direct action of GLP-1 on liver glucose metabolism or the glucagon effect of GLP-1 from our current methodology.

The liver effect of GLP-1 previously has been examined using *exogenous* GLP-1 infusion during pancreatic clamp, where plasma insulin and glucagon are not allowed to change and glycemia is maintained at basal concentrations or hyperglycemic levels [4, 5]. The findings from these two studies using different methods suggest that exogenous GLP-1 lowers endogenous glucose production in proportion to the increase in plasma glucose concentration; i.e., a greater *EGP* reducing effect of GLP-1 was observed during hyperglycemia compared to euglycemia (∽60 % [5] *versus* ∽20 % [4]).

Whether there is a glycemic threshold for liver effect of GLP-1 cannot be addressed by the current experimental design, but our data shows that *endogenous* GLP-1 has no influence on glucose counterregulatory response to hypoglycemia in fasting condition. Nonetheless, in our study, GLP-1R blockade enhanced prandial *EGP* response to hypoglycemia in GB subjects only but not in SG subjects or non-surgical controls despite a similar rise in glucagon concentration among the 3 groups. This suggests that variations in the effect of GLP-1 on liver glucose metabolism among GB *versus* SG or CN are independent of the plasma glucagon concentration. Also, consistent with previous observations [26-30], neither peripheral glucose disposal nor whole-body insulin sensitivity was affected by GLP-1R blockade. This novel observation indicates that, even at plasma glucose concentrations below fasting values, enhanced endogenous GLP-1 secretion and action after GB, can interfere with normal glucose counterregulatory response independent of insulin secretion or insulin sensitivity.

Despite sporadic case reports of postprandial hypoglycemia after SG [31], this condition, in our experience, is less prevalent, and likely to be of lesser severity than GB. The reason for this is unclear; however, a combination of more rapid surges of ingested nutrients and greater enteral beta-cell stimuli as well as altered prandial liver glucose metabolism after GB confer a higher risk for hypoglycemia compared to SG. Future studies are needed to directly examine the effect of GB *versus* SG on prandial liver glucose metabolism mediated by GLP-1 or glucagon.

A second major finding from this experiment is that alpha-cell but not beta-cell response to *endogenous* GLP-1 is preserved during hypoglycemia independently of bariatric surgery status. In patients with type 2 diabetes, the insulinotropic and glucagonostatic effects of *exogenous* GLP-1 infusion dissipate by normalization of the plasma glucose concentration [2], indicating that the pancreatic islet-cell effects of this peptide are glucose-dependent. In non-diabetic individuals with normal GI anatomy [32] or prior history of GB [33], however, acute administration of *exogenous* native GLP-1 or GLP-1R agonist results in greater insulin secretion during hyperinsulinemic hypoglycemic (2.7-3.0 mmol/l) clamp while glucagon inhibitory effect of these compounds disappears as glycemic concentration declines to 2.7-3.0 mmol/L. In contrast, we have observed that in non-diabetic subjects with or without bariatric surgery the *endogenous* GLP-1 has no effect on prandial insulin secretion during hypoglycemia (**∽**3.2 mmol/l) despite similar or greater prandial plasma GLP-1 concentrations achieved in the present study (120-250 pmol/l) (**Fig. 2A-B**) compared to those produced by *exogenous* GLP-1 infusion in the previous report (∽120 pmol/l) [32]. Further, the glucagon inhibitory effect of *endogenous* GLP-1, in our study, was preserved in prandial state under sub-basal glucose concentrations. Glucagon inhibitory effect of GLP-1 has been attributed to paracrine action of pancreatic produced GLP-1 on alpha-cell [34]. Therefore, the discrepancy between the present findings and those of previous reports can be explained by differences in methodology that highlights potential differences in glycemic threshold needed to turn off the insulinotropic or glucagonostatic properties of *endogenous versus exogenous* GLP-1 mediated by paracrine *versus* endocrine action of this peptide.

There are some limitations to our study that merit discussion. First, the sample size in each group of participants is relatively small reducing generalizability of these findings to a wider range of individuals after GB or SG. However, the groups were similar in gender, age, and BMI, and the surgical groups did not differ in the time and weight loss since surgery. Further, the effect of GLP-1R blockade on *EGP* response was consistent enough within GB subjects to result in highly significant differences among the 3 groups even with the modest number of studied subjects. Thus, despite a cross-sectional comparison, the results indicate that GB procedure has a significant effect on glucose counterregulatory response mediated by GLP-1. Also, antecedent hypoglycemia has been shown to alter glucose counter-regulation and *EGP* responses to future hypoglycemia [35] in non-diabetic humans. However, the counterregulatory adaptation should be similar during studies with and without Ex-9 infusion. Therefore, the reported *EGP* effect of GLP-1R blockade should not be affected by antecedent hypoglycemia in our study. Also, the rise in prandial glucose concentrations above the target level in 3 GB, 1 SG, and 1 CN could affect the *EGP* and glucagon responses. However, we found that the glycemic deviation after meal ingestion in this small subgroup is caused by a reduced action of insulin rather than alteration in *EGP* or *RaO* (**Supplementary Fig. 1**). Our target plasma insulin concentration is larger than observed in clinical conditions of hypoglycemia. Although, as previously demonstrated in humans [9, 36, 37] counterregulatory signals are still activated by hypoglycemia and can be evaluated at these levels of insulin.

In conclusion, we have shown that in obese individuals with and without prior bariatric surgery blockade of GLP-1R enhances prandial glucagon but not ISR response under hypoglycemic condition, suggesting that the insulinotropic activity but not glucagon inhibitor effect of *endogenous* GLP-1 is glucose-dependent. Although not affecting *Rd*, endogenous GLP-1 reduces (either directly or indirectly) prandial *EGP* response to hypoglycemia in post-GB individuals. The inhibitory effect of *endogenous* GLP-1 on *EGP* during insulin-induced hypoglycemia was consistent across our group of GB subjects. Taken together with our previous results which showed an enhanced prandial GLP-1-induced insulin secretion after GB [17-20] predisposing to hypoglycemia, these findings indicate that prandial GLP-1 also can interfere with counter-regulatory glucose responses that are important in prevention or correction of hypoglycemia in prandial state. Further studies are needed to examine the effectiveness of GLP-1R antagonists as therapeutic targets for improving the counterregulatory response to hypoglycemia in patients with GB but these observations provide a novel perspective on altered glucose metabolism following this procedure.

## METHODS

### Subjects

Eight patients with prior GB, seven patients with prior SG and five BMI-matched non-surgical controls of a cohort who were studied to address insulin clearance and glucose regulatory response to hypoglycemia [9, 23] were recruited in order of their presentation to the clinic or response to advertisement. None of the subjects had diabetes. Subjects with prior bariatric surgery had an average of 5 years since surgery and all subjects were weight stable for at least 3 months prior to enrollment. The Institutional Review Board of the University of Texas Health Science Center at San Antonio approved the protocol (HSC20180070H), and all subjects gave their written informed consent prior to participation.

### Peptides

Synthetic exendin-(9-39) (CS Bio, Menlo Park, California) was greater than 95 % pure, sterile, and free of pyrogens. Lyophilized peptide was prepared in 0.25 % human serum albumin on the day of study. The use of synthetic exendin-(9-39) is approved under the U.S. Food and Drug Administration Investigational New Drug 123,774.

### Experimental procedures

**S**ubjects were instructed to consume > 200 g of carbohydrate daily and not to engage in strenuous activity for three days prior to the experiments. Subjects reported to the Bartter Clinical Research Unit at Audie Murphy Hospital, South Texas Veteran Health Care System, in the morning after an overnight fast. Body composition was assessed using dual-energy X-ray absorptiometry (Hologic, Inc. Marlborough, Massachusetts, US), and waist circumference was measured. Intravenous catheters were placed in a hand vein and contralateral antecubital vein for blood sampling and infusion of glucose, insulin and exendin-(9-39), respectively. The hand was continuously kept warm with a heating pad.

### Mixed-meal test under hyperinsulinemic hypoglycemic conditions with and without exendin-(9-39) infusion (Fig. 1)

After fasting blood samples were drawn at –120 min, a prime-continuous infusion of [6,6-^2^H_2_]-glucose (28 μmol/kg prime and 0.28 μmol/kg/min constant) was initiated and continued for 2 hours, when the rate was reduced to half in anticipation of the insulin-mediated reduction in *EGP* for the remainder of the study [9]. At time point 0, a prime-continuous infusion of recombinant human insulin (Humulin 100 IU/ml) diluted in isotonic saline and mixed with 2 ml of the subjects’ blood was started and continued at 120 mU/min/m^2^ for the duration of the study [9]. Blood was sampled every 5-10 min and a variable infusion of 20 % glucose (1 % enriched with [6,6-^2^H_2_]-glucose) was infused to maintain plasma glucose concentration at a target of ∽3.2 mmol/l. At 60 minutes subjects received a prime-continuous infusion of exendin-(9-39) (Ex-9) (7500 pmol/kg prime and 750 pmol/kg/min constant) [16] or saline for the remainder of the study. At 120 minutes subjects ingested a 140 ml liquid meal containing 33 g whey protein, 12.7 g corn oil, and 14 g glucose mixed with 0.2 g [U-^13^C]-glucose over 10 minutes. This mixed-meal test replicates what is generally consumed by patients to treat acute hypoglycemia [25]. The plasma glucose concentration was maintained at the sub-basal glucose target (∽3.2 mmol/l) for 3 hours after meal ingestion. Blood samples were collected at timed intervals and stored in ice; plasma was separated within 60 minutes and stored at –80 °C until assayed.

### Assays

Plasma glucose was determined on-site using Analox GM9 Glucose Analyzer (Analox Instruments, Stourbridge, UK). Insulin (DIAsourve, Neuve, Belgium), C-peptide and glucagon (Millipore, Billerica, Massachusetts, US) were measured by commercial radioimmunoassay. The Millipore glucagon RIA kit has a sensitivity of ∽10 pmol/l and a cross-reactivity of <2 % with oxyntomodulin and glicentin [38]. GLP-1 and GIP were measured using commercial Multiplex ELISA (Millipore, Billerica, Massachusetts, US) according to manufacturer’s instructions. Tracer enrichment was measured using GC-MS, as previously described [39].

### Calculations

Fasting plasma glucose and hormone concentration represent the average of 3 samples drawn before the start of insulin. Insulin secretion rate (ISR) was derived from deconvolution of the plasma C-peptide concentration curve [23]. Rates of glucose disappearance (*Rd*), appearance of orally ingested glucose (*RaO*), and *EGP* were calculated from [6,6-^2^H_2_]-glucose and [U-^13^C]-glucose data using the Steele equation, as previously described [9, 16].

Prandial glucose fluxes and islet-cell and gut hormone responses were calculated as the incremental areas-under-the-curve (AUC) over pre-meal values using the trapezoidal rule. Whole-body insulin sensitivity was computed as the glucose clearance (*Rd*/glucose) divided by the corresponding plasma insulin concentration at fasting, steady-state period (110-120 min), and after meal ingestion (120-300 min) [40].

### Statistical analysis

Data are reported as mean ± standard error of mean. The effect of Ex-9 infusion *versus* saline infusion during clamp studies and the group effect (GB, SG, and CN), as well as their interaction on the outcomes of interest were analyzed using two-way ANOVA with repeated measures; Tukey honestly significant difference was used for *post hoc* analysis to compare differences among 3 groups noted earlier. The variables of interest at baseline were compared with χ2 test or ANOVA among the groups. Statistical analyses were done using SPSS 28 (SPSS Inc., Chicago, IL). The STROBE cross-sectional reporting guidelines were used [41].

## Data Availability

All data produced in the present study are available upon reasonable request to the authors.

## Abbreviations

CN: non-surgical controls
*EGP*: endogenous glucose production
Ex-9: exendin-(9-39)
GB: gastric bypass
GIR: glucose infusion rate
GLP-1: glucagon-like peptide 1
ICR: insulin clearance
ISR: insulin secretion rate
*Ra*: rate of glucose appearance
*Rd*: rate of glucose disappearance
*RaO*: rate of oral glucose appearance
SG: sleeve gastrectomy

## Author contributions

H.H. analyzed and interpreted the data and drafted the manuscript. A.G., S.P., R.P., and R.D. reviewed and revised the manuscript for important intellectual content. M.S. is the guarantor of this work and, as such, takes responsibility for the work as a whole, including the study design, access to data, and the decision to submit and publish the manuscript. All authors approved the final version of the manuscript.

## LEGENDS FOR FIGURES

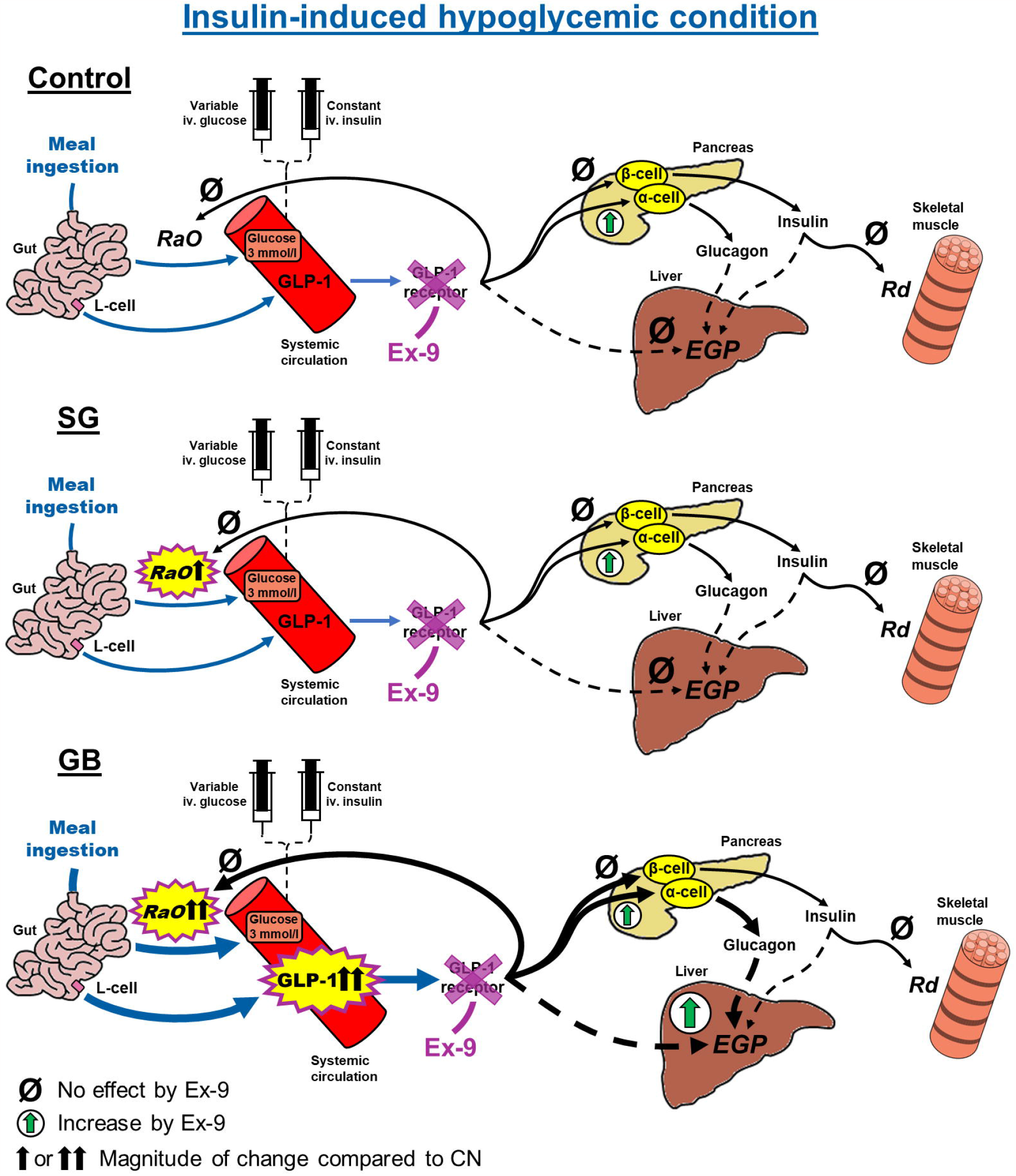

