## Supplemental Material for "Endogenous glucagon-like peptide 1 diminishes prandial glucose counterregulatory response to hypoglycemia after gastric bypass surgery"

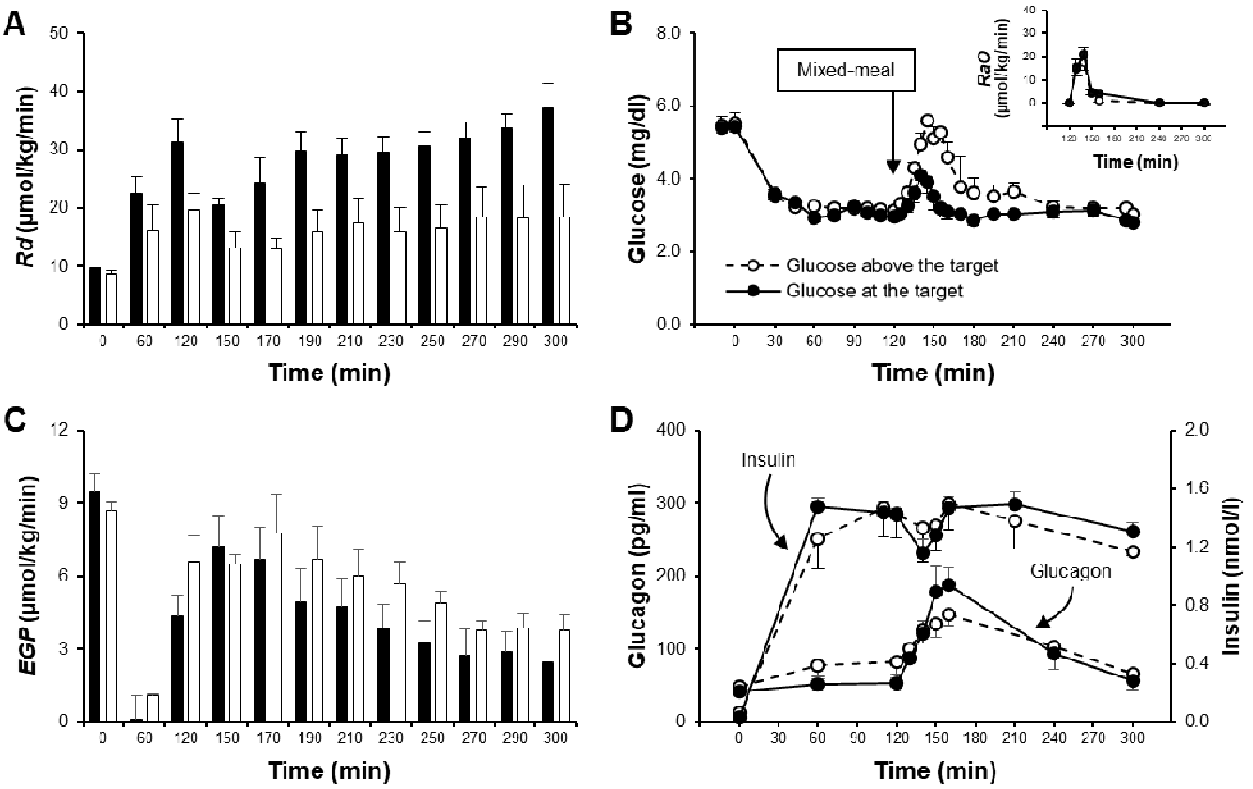


**Supplementary Figure 1.** ***Rd*** **(A), plasma glucose and** ***RaO*** **(B),** ***EGP*** **(C) and islet-cell hormone concentrations (D) in GB-treated subjects who had their plasma glucose at (n = 5, black bars and solid lines) or above the target glucose (n = 3, white bars and dashed lines) of ~3.2 mmol/l after meal ingestion.**

**Supplementary Table 1. Heart rate and blood pressure responses to insulin-induced hypoglycemia and mixed-meal test during saline and exendin-(9-39) infusion in GB, SG and CN groups.**

| | | Saline | | | Ex-9 | | | Two-way ANOVA | | |
|---|---|---|---|---|---|---|---|---|---|---|
| Parameter | Time (min) | **GB** | **SG** | **CN** | **GB** | **SG** | **CN** | **T** | **G** | **I** |
| Heart rate (bpm) | Basal | 67.5 ± 3.3 | 64.9 ± 4.5 | 74.1 ± 4.8 | 62.7 ± 3.1 | 61.8 ± 4.7 | 77.9 ± 6.1 | 0.328 | 0.125 | 0.053 |
| | Pre-meal | 74.5 ± 5.4 | 68.9 ± 4.2 | 85.1 ± 2.9* | 72.9 ± 6.1 | 68.3 ± 3.4 | 86.3 ± 5.9* | 0.918 | **0.043** | 0.950 |
| | Prandial 1hr | 83.8 ± 4.8 | 75.6 ± 5.1 | 84.6 ± 4.6 | 84.9 ± 5.6 | 76.1 ± 5.6 | 87.0 ± 6.2 | 0.699 | 0.288 | 0.976 |
| | Prandial 3hrs | 80.4 ± 5.0 | 72.6 ± 4.1 | 83.8 ± 3.7 | 76.7 ± 5.6 | 70.7 ± 4.1 | 85.0 ± 6.6 | 0.680 | 0.148 | 0.852 |
| Systolic BP (mmHg) | Basal | 115.8 ± 5.1 | 121.8 ± 3.9 | 126.2 ± 4.2 | 111.1 ± 4.7 | 122.5 ± 5.1 | 123.3 ± 3.5 | 0.121 | 0.207 | 0.248 |
| | Pre-meal | 117.0 ± 6.6 | 119.7 ± 4.5 | 126.4 ± 3.9 | 111.9 ± 5.4 | 119.6 ± 5.2 | 121.7 ± 2.6 | 0.224 | 0.415 | 0.679 |
| | Prandial 1hr | 122.7 ± 7.1 | 116.7 ± 5.9 | 121.8 ± 3.9 | 113.4 ± 4.5 | 120.2 ± 4.4 | 119.9 ± 4.5 | 0.289 | 0.930 | 0.081 |
| | Prandial 3hrs | 118.5 ± 6.6 | 114.8 ± 4.7 | 121.9 ± 4.7 | 108.5 ± 4.7 | 119.1 ± 4.1 | 114.4 ± 7.8 | 0.055 | 0.803 | **0.023** |
| Diastolic BP (mmHg) | Basal | 72.2 ± 3.3 | 69.6 ± 3.4 | 69.5 ± 5.5 | 67.7 ± 3.4 | 70.8 ± 3.5 | 77.0 ± 2.2 | 0.243 | 0.789 | **0.002** |
| | Pre-meal | 68.9 ± 4.1 | 62.1 ± 2.9 | 69.9 ± 5.0 | 63.8 ± 4.0 | 64.6 ± 3.3 | 71.8 ± 3.5 | 0.910 | 0.380 | 0.193 |
| | Prandial 1hr | 68.7 ± 4.4 | 59.3 ± 4.0 | 65.5 ± 5.4 | 62.0 ± 3.2 | 64.9 ± 3.1 | 68.3 ± 3.8 | 0.734 | 0.681 | **0.016** |
| | Prandial 3hrs | 67.2 ± 4.3 | 59.0 ± 3.3 | 65.2 ± 5.7 | 60.8 ± 3.2 | 63.9 ± 3.2 | 68.7 ± 3.3 | 0.662 | 0.58 | **0.004** |

Data are presented as mean ± SEM. Statistical effect *P* values (treatment: saline *versus* Ex-9 [T]; group status: GB, SG, or CN [G]; and their interaction [I]) are provided in the 3 columns on the right. †$P < 0.05$ compared with GB.

BP, blood pressure; CN, non-surgical controls; Ex-9, exendin-(9-39); GB, subjects with prior gastric bypass surgery; SG, subjects with prior sleeve gastrectomy.
